# A Quick Displacement of the SARS-CoV-2 variant Delta with Omicron: Unprecedented Spike in COVID-19 Cases Associated with Fewer Admissions and Comparable Upper Respiratory Viral Loads

**DOI:** 10.1101/2022.01.26.22269927

**Authors:** Amary Fall, Raghda E. Eldesouki, Jaiprasath Sachithanandham, C. Paul Morris, Julie M. Norton, David C. Gaston, Michael Forman, Omar Abdullah, Nicholas Gallagher, Maggie Li, Nicholas J. Swanson, Andrew Pekosz, Eili Y. Klein, Heba H. Mostafa

## Abstract

**Background:** The increase in SARS-CoV-2 infections in December 2021 in the United States was driven primarily by the Omicron variant which largely displaced the Delta over a three week span. Outcomes from infection with the Omicron remain uncertain. We evaluate whether clinical outcomes and viral loads differ between Delta and Omicron infections during the period when both variants were co-circulating.

**Methods:** Remnant clinical specimens from patients that tested positive for SARS-CoV-2 after standard of care testing between the last week of November and the end of December 2021were used for whole viral genome sequencing. Cycle threshold values (Ct) for viral RNA, the presence of infectious virus, and levels of respiratory IgG were measured, and clinical outcomes were obtained. Differences in each measure were compared between variants stratified by vaccination status.

**Results:** The Omicron variant displaced the Delta during the study period and constituted 95% of the circulating lineages by the end of December 2021. Patients with Omicron infections (N= 1121) were more likely to be vaccinated compared to patients with Delta (N = 910), but were less likely to be admitted, require ICU level care, or succumb to infection regardless of vaccination status. There was no significant difference in Ct values based on the lineage regardless of the vaccination status. Recovery of infectious virus in cell culture was reduced in boosted patients compared to fully vaccinated without a booster and unvaccinated when infected with the Delta lineage. However, in patients with Omicron infections, recovery of infectious virus was not affected by vaccination.

**Conclusions:** Omicron infections of vaccinated individuals are expected, yet admissions are less frequent. Admitted patients might develop severe disease comparable to Delta. Efforts for reducing the Omicron transmission are required as even though the admission risk is lower, the numbers of infections continue to be high.

**Research in context Evidence before this study:** The unprecedented increase in COVID-19 cases in the month of December 2021, associated with the displacement of the Delta variant with the Omicron, triggered a lot of concerns. An understanding of the disease severity associated with infections with Omicron is essential as well as the virological determinants that contributed to its widespread predominance. We searched PubMed for articles published up to January 23, 2022, using the search terms (“Omicron”) AND (“Disease severity”) as well as (“Omicron”) AND (“Viral load”) And/ or (“Cell culture”). Our search yielded 3 main studies that directly assessed the omicron’s clinical severity in South Africa, its infectious viral load compared to Delta, and the dynamics of viral RNA shedding. In South Africa, compared to Delta, Omicron infected patients showed a significant reduction in severe disease. In this study, Omicron and non-Omicron variants were characterized based on S gene target failure using the TaqPath COVID-19 PCR (Thermo Fisher Scientific). In the study from Switzerland that assessed the infectious viral load in Omicron versus Delta, the authors analyzed only 18 Omicron samples that were all from vaccinated individuals to show that compared to Delta, Omicron had equivalent infectious viral titers. The third study that assessed the Omicron viral dynamics showed that the peak viral RNA in Omicron infections is lower than Delta. No published studies assessed the clinical discrepancies of Omicron and Delta infected patients from the US, nor comprehensively assessed, by viral load and cell culture studies, the characteristics of both variants stratified by vaccination status.

**Added value of this study:** To the best of our knowledge, this is the only study to date to compare the clinical characteristics and outcomes after infection with the Omicron variant compared to Delta in the US using variants characterized by whole genome sequencing and a selective time frame when both variant co-circulated. It is also the first study to stratify the analysis based on the vaccination status and to compare fully vaccinated patients who didn’t receive a booster vaccination to patients who received a booster vaccination. In addition, we provide a unique viral RNA and infectious virus load analyses to compare Delta and Omicron samples from unvaccinated, fully vaccinated, and patients with booster vaccination.

**Implications of all the available evidence:** Omicron associated with a significant increase in infections in fully and booster vaccinated individuals but with less admissions and ICU level care. Admitted patients showed similar requirements for supplemental oxygen and ICU level care when compared to Delta admitted patients. Viral loads were similar in samples from Omicron and Delta infected patients regardless of the vaccination status. The recovery of infectious virus on cell culture was reduced in samples from patients infected with Delta who received a booster dose, but this was not the case with Omicron. The recovery of infectious virus was equivalent in Omicron infected unvaccinated, fully vaccinated, and samples from patients who received booster vaccination.

**Funding:** NIH/NIAID Center of Excellence in Influenza Research and Surveillance contract HHS N2772201400007C, Johns Hopkins University, Maryland department of health, Centers for Disease Control and Prevention contract 75D30121C11061.

## Introduction

The SARS-CoV-2 Omicron variant was first identified in South Africa and reported to the World Health Organization (WHO) on November 24, 2021 (1). A large number of mutations and amino acid changes were noted across the Omicron genome, 15 of which are within the receptor binding domain (RBD) of the spike (S) protein (2). Some of the Omicron RBD characterized mutations raised concerns of a significant impact on the transmissibility, immunity secondary to vaccination or prior infection, and efficacy of therapeutic monoclonal antibodies. Hence, the WHO designated the Omicron as a variant of concern (VOC) on November 26, 2021 (3, 4). Initial reports from South Africa (5) and the UK (6) suggested that the Omicron variant may be more transmissible but cause less severe infection. However, the US population differs from both the South African and UK populations in significant ways; most importantly, the percentage of the population that is vaccinated is significantly lower in the US than the UK and prior infection is significantly lower in the US compared to South Africa. Thus, significant questions remain as to the impact Omicron will have on the US population. Most importantly, how might outcomes, particularly hospitalization, differ between patients infected with Omicron compared to variants such as Delta. Additionally, there remains uncertainty as to whether enhanced evasion of pre-existing immunity or some other biological mechanisms drive the higher rate of Omicron transmission.

The Omicron variant was first identified in Maryland in the last week of November 2021, and became predominant in a matter of 3 weeks. In this study, we evaluated the biological differences between viral variants collected as a part of routine clinical care between November 22^nd^ and December 31^st^ in the Johns Hopkins Health System, as well as the clinical outcomes in patients infected with Omicron and Delta. We focused on this time frame as it witnessed the detection of the first Omicron case in our system in the last week of November, and the switch between Delta predominance in the beginning of December to Omicron predominance at the end of December 2021. We provide a comparison of clinical, demographic and virological load between Delta and Omicron infected individuals and stratify our results based on vaccination status.

## Methods

### Ethical considerations and Data availability

Research was conducted under Johns Hopkins IRB protocol IRB00221396 with a waiver of consent. Remnant clinical specimens from patients that tested positive for SARS-CoV-2 after standard of care testing were used for whole genome sequencing. Whole genomes were made publicly available at GISAID.

### Specimens and Patient Data

Nasopharyngeal or lateral mid-turbinate nasal swabs after standard of care diagnostic or screening testing were collected and used for genome sequencing. At Johns Hopkins Medical System, SARS-CoV-2 clinical testing is performed for inpatients and outpatients (five acute care hospitals and more than 40 ambulatory care offices) as well as standard of care screening particularly prior to scheduled surgeries.

Molecular assays used include primarily the NeuMoDx SARS-CoV-2 (Qiagen), Cobas SARS-CoV-2 (Roche), Xpert Xpress SARS-CoV-2/Flu/RSV (Cepheid), in addition to the RealStar® SARS-CoV-2 RT- PCR (Altona Diagnostics), ePlex Respiratory Pathogen Panel 2 (Roche), Aptima SARS-CoV-2 (Hologic), and Accula SARS-CoV-2 assays (ThermoFisher Scientific) (7-10). Each sample in our cohort represents a unique patient. Table 1 summarizes the numbers of charts and samples used for each part of the study.

**Table 1.** Charts and samples used for the study.

|  | Omicron |  |  |  | Delta |  |  |  |
| --- | --- | --- | --- | --- | --- | --- | --- | --- |
| Total Patients | 1121 |  |  |  | 910 |  |  |  |
|  | Fully Vaccinated | With booster dose | Unvaccinated | Partially vaccinated | Fully Vaccinated | With booster dose | Unvaccinated | Partially vaccinated |
| Charts | 461 | 134 | 488 | 38 | 282 | 83 | 518 | 27 |
| Ct analysis | 184 | 51 | 170 | 14 | 195 | 45 | 411 | 9 |
| Patients with known days from the onset of symptoms | 159 | 51 | 135 | 12 | 112 | 16 | 225 | 5 |
| IgG | 152 | 43 | 0 | 11 | 183 | 43 | 0 | 7 |
| Cell culture (total) | 55 | 76 | 83 | 5 | 52 | 45 | 56 | 0 |
| Cell culture (Ct ≤ 20) | 50 | 41 | 56 |  | 49 | 27 | 44 |  |

### Clinical data analysis

Clinical and vaccination data for patients whose samples were characterized by whole genome sequencing was bulk extracted as previously detailed in (11). Repeat tests from the same patient were excluded as were results with uncharacterized genomes due to insufficient quality (n=347). The first Omicron infection was collected from a patient in the Johns Hopkins Medical System during the last week of November, 2022. Notably, samples are collected randomly from the whole system for whole genome sequencing for surveillance. A total of 1121 Omicron and 910 Delta infected patients diagnosed between November 22nd 2021 and December 31st 2021 were included in the study. COVID admission relatedness was determined based on presenting complaints, admission diagnoses, reason for testing, and timing of testing. Patients admitted with no COVID-related complaints and no COVID-related diagnoses that were tested after admission as part of an asymptomatic screening protocol that is administered to all admitted patients were considered non-COVID-related admissions. Additionally, patients who developed symptoms after admission or who tested positive on regular asymptomatic surveillance of inpatients were also considered non-COVID-related admissions. In our vaccinated patients’ population, 68.6% received Pfizer/BioNTech, followed by the Moderna mRNA-1273 (26.6%), then the J&J/Janssen COVID-19 vaccines (4.8%). Full vaccination was based on the CDC definition of positive test results more than 14 days post the second shot for pfizer/BioNTech BNT162b2 and Moderna mRNA-1273 or 14 days after the J&J/Janssen.

### Ct value analysis

To ensure comparable Ct values for viral load analyses, samples were retested with the PerkinElmers SARS-CoV-2 kit (https://www.fda.gov/media/136410/download) and Ct values of the N gene were used for comparisons.

### Amplicon based Sequencing

Specimens preparation, extractions, and sequencing were performed as described previously (12, 13). Library preparation for this cohort was performed using the NEBNext® ARTIC SARS-CoV-2 Companion Kit (VarSkip Short SARS-CoV-2 # E7660-L). Sequencing was performed using the Nanopore GridION and reads were basecalled with MinKNOW, and demultiplexed with guppybarcoder that required barcodes at both ends. Alignment and variant calling were performed with the artic- ncov2019 medaka protocol with thresholds set to a minimum of 90% coverage and 100 mean depth.

Query mutations were manually confirmed with Integrated Genomics viewer (IGV) (Version 2.8.10), clades were determined using Nextclade beta v 0.12.0 (clades.nextstrain.org), and lineages were determined with Pangolin COVID-19 lineage Assigner (COG-UK (cog-uk.io)).

### ELISA

Respiratory samples were tested, undiluted, with the EUROIMMUN Anti-SARS-CoV-2 ELISA (IgG) following the package insert (https://www.fda.gov/media/137609/download) as we described previously (11). The assay detects antibodies to the SARS-CoV-2 S1 domain of the spike protein. The value 1.1 was used as a cut off for positives.

### Cell culture

Vero-TMPRSS2 cells were cultured and infected with aliquots of swab specimens as previously described for VeroE6 cells (14). Cultures were incubated for 6 days and SARS-CoV-2 cytopathic effect (CPE) was confirmed by reverse transcriptase PCR.

### Statistical analysis

Statistical analyses were conducted using GraphPad prism. Chi-square and Fisher Exact tests were used for categorical variable comparisons and t-test and one-way ANOVA were used for comparing continuous independent variables.

## Results

### SARS-CoV-2 positivity and variants trends November- December 2021

The SARS-CoV-2 positivity rate increased markedly in December 2021 (25.5%, Figure 1A). The large increase in SARS-CoV-2 positivity in the month of December was the highest recorded since the beginning of the pandemic (Figure S1 and (15)). The spike in SARS-CoV-2 positivity was particularly evident during the last two weeks of December for both symptomatic and asymptomatic patients (Figure 1A). The increase in the positivity correlated with an increase in the detection of the Omicron variant, that went from less than 1% of sequenced strains in the beginning of December to the dominant variant in less than 3 weeks (Figure 1B and C). Table 2 shows the total numbers tested and sequenced and the detailed positivity per respiratory target.

**Figure 1.**
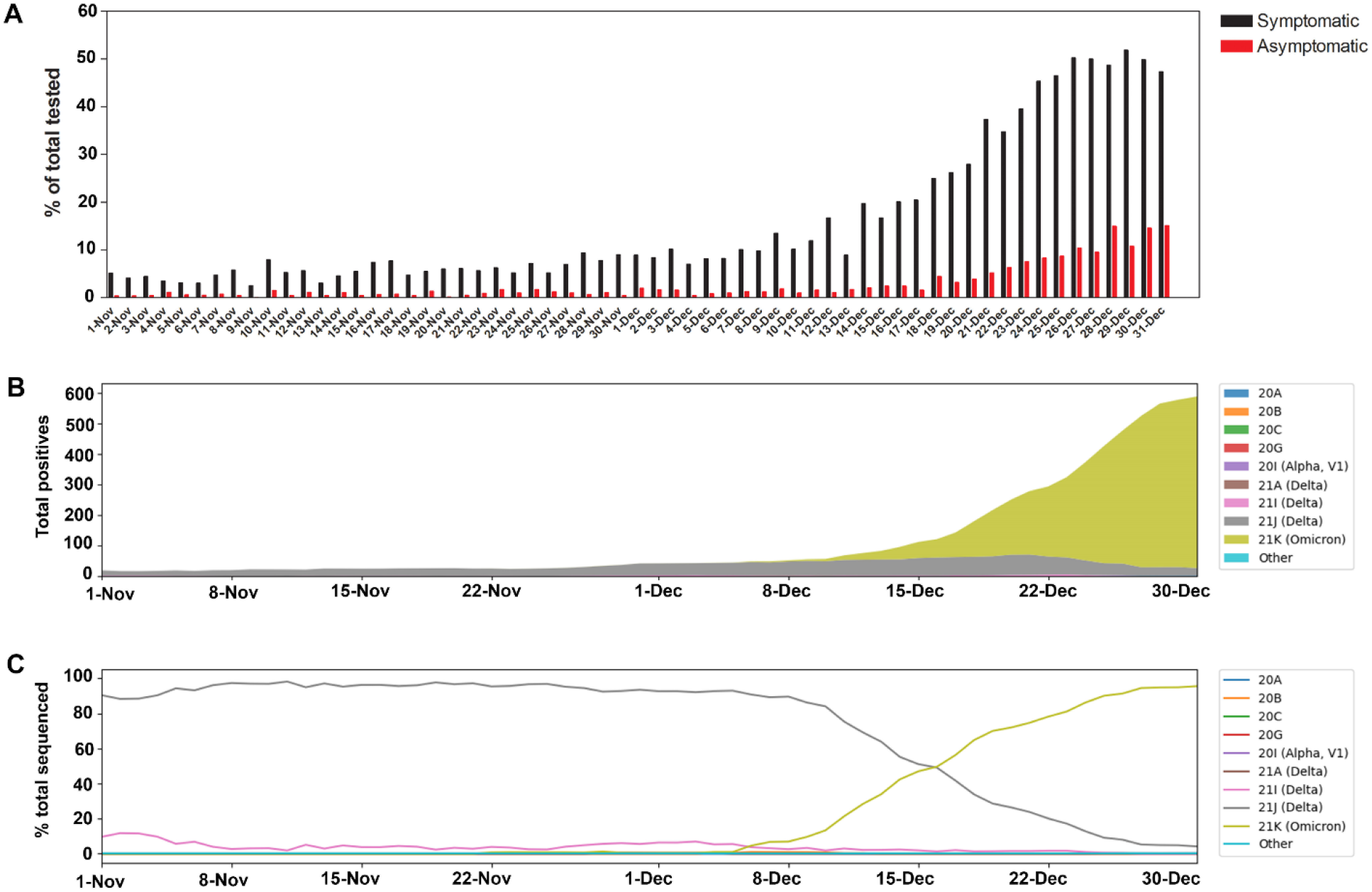
SARS-CoV-2 positivity and variants trends November- December 2021. A) SARS-CoV-2 daily positivity rates between November and December 2021 for both symptomatic and asymptomatic testing. A) SARS-CoV-2 clade distribution between November and December 2021 relative to the 7 day rolling average positives from Johns Hopkins system. C) Percent SARS-CoV-2 clades in November and December 2021.

**Table 2.** SARS-CoV-2 testing and positivity in November and December 2021 and total sequenced. SARS-CoV-2 testing is performed by different molecular assays as detailed in the methods section and the total tested reflects testing by all assays. Sequence counts were up to the time of writing this manuscript.

|  | November | December | November | December |
| --- | --- | --- | --- | --- |
| **SARS-CoV-2 (All testing)** | **726** | **7078** | **2.67** | **18.65** |
| **SARS-CoV-2 (Symptomatic only)** | **592** | **6236** | **5.70** | **25.50** |
| **SARS-CoV-2 whole genome sequencing** |  |  | **%** | **%** |
| **Total sequenced** | **552** | **2179** | **76.03** | **30.79** |
| **21I (Delta)** | **24** | **36** | **4.35** | **1.65** |
| **21J (Delta)** | **468** | **791** | **84.78** | **36.30** |
| **21K (Omicron)** | **1** | **1208** | **0.18** | **55.44** |
| **Other or low QC** | **59** | **144** | **10.69** | **6.61** |

### Patient characteristics and infection outcomes in Omicron and Delta infections

A total of 7,353 samples tested positive for SARS-CoV-2 of a total 45,856 tested in the Johns Hopkins Laboratories between November 22nd 2021 and December 31st 2021. Of these, 2,378 were randomly selected for whole genome sequencing. After excluding repeat tests in patients and results that were unable to be characterized due to low quality, a total of 2,031 patients, 1121 Omicron and 910 Delta, were included in the study. Patients infected with the Omicron variant were younger (median age 32 years vs 35, p < 0.001, Table 3) and significantly more likely to be vaccinated than patients with Delta infections (Table 3). However, compared to patients with Delta, patients with Omicron were significantly less likely to be admitted (3 % vs 13.8 %, p < 0.00001), require ICU level care (0.5% vs 3.5%, p < 0.00001), or expire (0.1 vs 1.1, p = 0.004, Table 3).

**Table 3.**
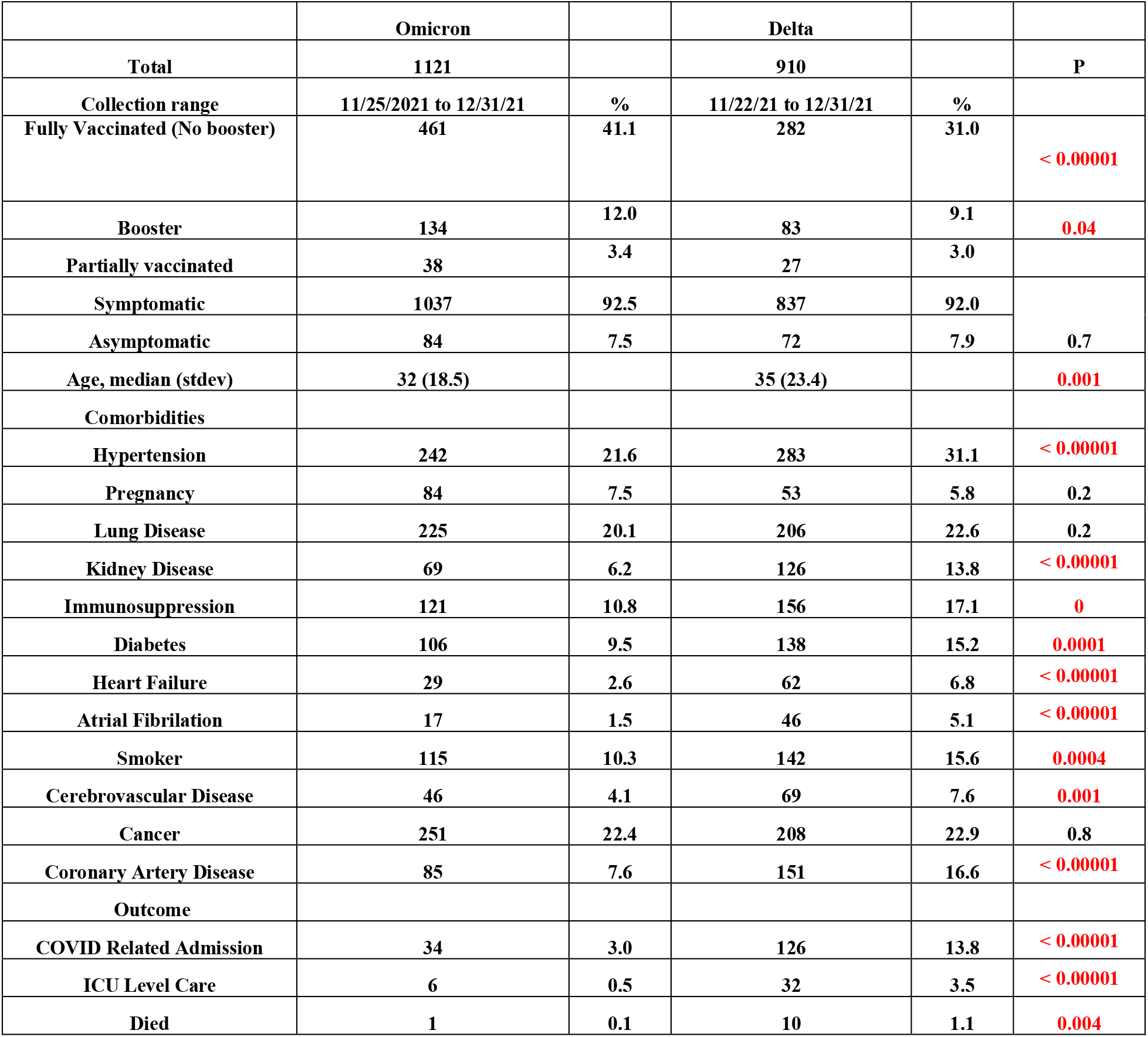
Clinical and metadata of the Omicron and Delta infected patients. Statistics for ages were calculated by *t* test and all other statistics were calculated by Chi-squared test.

|  | Omicron |  | Delta |  |  |
| --- | --- | --- | --- | --- | --- |
| Total | 1121 |  | 910 |  | P |
| Collection range | 11/25/2021 to 12/31/21 | % | 11/22/21 to 12/31/21 | % |  |
| Fully Vaccinated (No booster) | 461 | 41.1 | 282 | 31.0 | < 0.00001 |
| Booster | 134 | 12.0 | 83 | 9.1 | 0.04 |
| Partially vaccinated | 38 | 3.4 | 27 | 3.0 |  |
| Symptomatic | 1037 | 92.5 | 837 | 92.0 | 0.7 |
| Asymptomatic | 84 | 7.5 | 72 | 7.9 |  |
| Age, median (stdev) | 32 (18.5) |  | 35 (23.4) |  | 0.001 |
| Comorbidities |  |  |  |  |  |
| Hypertension | 242 | 21.6 | 283 | 31.1 | < 0.00001 |
| Pregnancy | 84 | 7.5 | 53 | 5.8 | 0.2 |
| Lung Disease | 225 | 20.1 | 206 | 22.6 | 0.2 |
| Kidney Disease | 69 | 6.2 | 126 | 13.8 | < 0.00001 |
| Immunosuppression | 121 | 10.8 | 156 | 17.1 | 0 |
| Diabetes | 106 | 9.5 | 138 | 15.2 | 0.0001 |
| Heart Failure | 29 | 2.6 | 62 | 6.8 | < 0.00001 |
| Atrial Fibrillation | 17 | 1.5 | 46 | 5.1 | < 0.00001 |
| Smoker | 115 | 10.3 | 142 | 15.6 | 0.0004 |
| Cerebrovascular Disease | 46 | 4.1 | 69 | 7.6 | 0.001 |
| Cancer | 251 | 22.4 | 208 | 22.9 | 0.8 |
| Coronary Artery Disease | 85 | 7.6 | 151 | 16.6 | < 0.00001 |
| Outcome |  |  |  |  |  |
| COVID Related Admission | 34 | 3.0 | 126 | 13.8 | < 0.00001 |
| ICU Level Care | 6 | 0.5 | 32 | 3.5 | < 0.00001 |
| Died | 1 | 0.1 | 10 | 1.1 | 0.004 |

In the vaccinated population, patients with either Omicron or Delta were likely to be older, though this was only significant in the Omicron group (Table 4). No significant differences in comorbidities or disease severity was noted when all fully vaccinated were compared to unvaccinated for both Delta and Omicron groups (Table 4). However, in fully vaccinated patients who had received a booster, patients with Delta infections were significantly more likely to have comorbidities including immunosuppression and lung disease and were more likely to be admitted than patients with Omicron (Table S1). While patients admitted with Omicron infections were more commonly fully vaccinated (58.8 % vs 23.8 %, p = 0.0001), we didn’t notice a significant difference in the need for supplemental oxygen, ICU level care, or mortality (Table 5).

**Table 4.** Clinical and metadata of Delta and Omicron vaccinated and unvaccinated patients. Statistics for ages were calculated by t test and all other statistics were calculated by Chi-squared test. stdev; standard deviation.

|  | Omicron |  |  |  |  | Delta |  |  |  |  |
| --- | --- | --- | --- | --- | --- | --- | --- | --- | --- | --- |
|  | Fully vaccinated including boosted |  | Unvaccinated |  |  | Fully vaccinated including boosted |  | Unvaccinated |  |  |
| Total | 595 | % | 488 | % | p | 365 | % | 518 | % | p |
| Symptomatic | 547 | 91.93 | 454 | 93.03 | 0.60 | 329 | 90.14 | 385 | 74.32 | 0.3 |
| Asymptomatic | 48 | 8.07 | 34 | 6.97 |  | 36 | 9.86 | 32 | 6.18 |  |
| Age, median (stdev) | 33 (18.2) |  | 31.8 (18.8) |  | 0.02 | 37 (23.2) |  | 34 (23.6) |  | 0.09 |
| Comorbidities |  |  |  |  |  |  |  |  |  |  |
| Hypertension | 127 | 21.34 | 109 | 22.34 | 0.70 | 126 | 34.52 | 149 | 28.76 | 0.07 |
| Pregnancy | 50 | 8.40 | 30 | 6.15 | 0.20 | 24 | 6.58 | 27 | 5.21 | 0.50 |
| Lung Disease | 107 | 17.98 | 111 | 22.75 | 0.06 | 84 | 23.01 | 115 | 22.20 | 0.80 |
| Kidney Disease | 36 | 6.05 | 30 | 6.15 | 1.00 | 50 | 13.70 | 72 | 13.90 | 1.00 |
| Immunosuppression | 66 | 11.09 | 51 | 10.45 | 0.80 | 65 | 17.81 | 86 | 16.60 | 0.60 |
| Diabetes | 60 | 10.08 | 43 | 8.81 | 0.50 | 60 | 16.44 | 74 | 14.29 | 0.40 |
| Heart Failure | 15 | 2.52 | 13 | 2.66 | 1.00 | 30 | 8.22 | 30 | 5.79 | 0.20 |
| Atrial Fibrillation | 9 | 1.51 | 8 | 1.64 | 1.00 | 18 | 4.93 | 27 | 5.21 | 0.90 |
| Smoker | 64 | 10.76 | 45 | 9.22 | 0.40 | 53 | 14.52 | 84 | 16.22 | 0.50 |
| Cerebrovascular Disease | 24 | 4.03 | 20 | 4.10 | 1.00 | 26 | 7.12 | 41 | 7.92 | 0.70 |
| Cancer | 142 | 23.87 | 103 | 21.11 | 0.30 | 91 | 24.93 | 112 | 21.62 | 0.30 |
| Coronary Artery Disease | 51 | 8.57 | 31 | 6.35 | 0.20 | 60 | 16.44 | 88 | 16.99 | 0.80 |
| Outcome |  |  |  |  |  |  |  |  |  |  |
| COVID Related Admission | 23 | 3.87 | 9 | 1.84 | 0.07 | 43 | 11.78 | 80 | 15.44 | 0.13 |
| ICU Level Care | 3 | 0.50 | 2 | 0.41 | 1.00 | 8 | 2.19 | 24 | 4.63 | 0.07 |
| Died | 0 | 0.00 | 1 | 0.20 | 0.40 | 4 | 1.10 | 6 | 1.16 | 1.00 |

**Table 5.** Clinical and metadata of Delta and Omicron admitted patients. Statistics for ages were calculated by t test and all other statistics were calculated by Chi-squared test.

|  | Omicron admissions |  | Delta admissions |  |  |
| --- | --- | --- | --- | --- | --- |
| Total | 34 | % | 126 | % | p value |
| Fully vaccinated | 20 | 58.8 | 30 | 23.8 | 0.0001 |
| Booster | 3 | 8.8 | 13 | 10.3 | 1 |
| Median age (stdev) | 48 (25.8) |  | 58 (21.1) |  | 0.130 |
| Comorbidities |  |  |  |  |  |
| Hypertension | 17 | 50.0 | 82 | 65.1 | 0.11 |
| Pregnancy | 4 | 11.8 | 4 | 3.2 | 0.06 |
| Lung Disease | 11 | 32.4 | 38 | 30.2 | 0.8 |
| Kidney Disease | 18 | 52.9 | 61 | 48.4 | 0.7 |
| Immunosuppression | 20 | 58.8 | 59 | 46.8 | 0.24 |
| Diabetes | 13 | 38.2 | 51 | 40.5 | 0.84 |
| Heart Failure | 10 | 29.4 | 25 | 19.8 | 0.24 |
| Atrial Fibrillation | 7 | 20.6 | 27 | 21.4 | 1 |
| Smoker | 8 | 23.5 | 32 | 25.4 | 1 |
| Cerebrovascular Disease | 7 | 20.6 | 26 | 20.6 | 1 |
| Cancer | 12 | 35.3 | 46 | 36.5 | 1 |
| Coronary Artery Disease | 15 | 44.1 | 64 | 50.8 | 0.56 |
| Outcome |  |  |  |  |  |
| Supplemental O2 | 23 | 67.6 | 92 | 73.0 | 0.5 |
| ICU Level Care | 6 | 17.6 | 32 | 25.4 | 0.5 |
| Died | 1 | 2.9 | 10 | 7.9 | 0.5 |

### Omicron and Delta variants cycle threshold (Ct) values in upper respiratory samples

To determine if the Ct values in upper respiratory samples were different in Omicron versus Delta infected individuals, we compared the Ct values collected for all the groups regardless of the days to the onset of symptoms or the status of the patients being symptomatic or asymptomatic (Omicron = 419, Delta N = 660). No difference in the mean or median Ct values were notable (Figure 2A). No differences were noted when Ct values were compared between vaccinated and unvaccinated patients from both groups as well (Omicron vaccinated = 235, unvaccinated = 170, Delta vaccinated = 240, unvaccinated = 411, Figure 2A). When the Ct analysis was correlated to the days from the onset of symptoms for symptomatic patients, no significant differences were detected between Omicron and Delta fully vaccinated, boosted, or unvaccinated (Figure 2B and C and Figure S2). We conclude that there were no significant differences in viral RNA loads between Omicron and Delta infected individuals.

**Figure 2.**
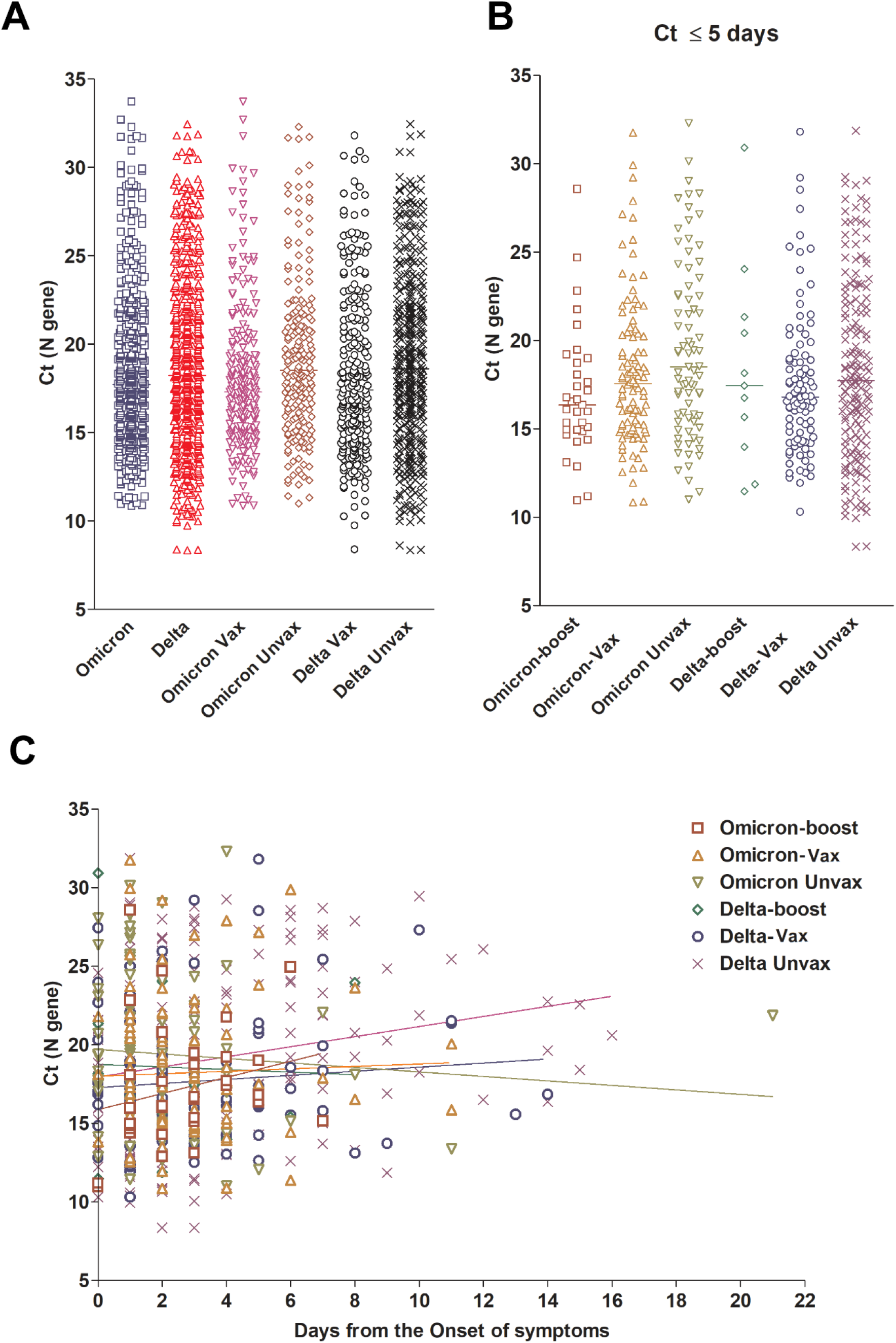
Omicron and Delta variants cycle threshold (Ct) values in upper respiratory samples. A) Ct values of Omicron and Delta from all samples with available Ct values (N gene) stratified by vaccination status. B) Ct values of Omicron and Delta from samples collected 5 days or less from the onset of symptoms. For this analysis, samples from asymptomatic patients were not included. C) Ct values of Omicron and Delta variants broken down by vaccination status and associated with days after the onset of symptoms. Vax, fully vaccinated patients who didn’t receive a booster dose (panel A only, Vax includes boosted patients); Unvax, unvaccinated; boost, patients with booster dose.

### Recovery of infectious virus in Omicron versus Delta groups

To assess the recovery of infectious virus from upper respiratory tract specimens of individuals infected with Omicron versus Delta variants, samples from 219 Omicron (N, Table 1) and 153 Delta (N, Table 1) infections were used to inoculate Vero-TMPRSS2 cells. Recovery of infectious virus (positive cytopathic effects; CPE) was noted from more specimens from the Delta group as compared to the Omicron group (Delta 78%, Omicron 61%; Figure 3A, p = 0.0006). Specimens from the boosted Delta group showed significant reduction in the recovery of infectious virus as compared to the fully vaccinated or the unvaccinated Delta groups (62% vs 87 and 82 %, p = 0.009 and 0.04, Figure 3A). This was not the case in the Omicron groups which had no statistically significant differences in infectious virus recovery between the boosted, fully vaccinated and unvaccinated groups (55%, 69%, and 60%, Figure 3A). A significant increase in the recovery of infectious virus from specimens of patients infected with the Delta variant as compared to the Omicron was noted for both fully vaccinated (87% vs 69%, p < 0.04) and unvaccinated (82% vs 60%, p = 0.008) groups (Figure 3A). Consistent with our previously published reports (Figure 3C and (11, 14)), and since lower Ct values have been associated with positive CPE, we compared samples with Ct values less than 20 for both Delta and Omicron groups (N; Table 1). Our analysis showed that the Delta infection was associated a significant increase in samples with positive CPE compared to Omicron (87% vs 70%, p = 0.001) however, no significant differences were seen between boosted, fully vaccinated, or unvaccinated groups (Figure 3B). Taken together, Omicron infections had lower numbers of samples with infectious virus when compared to Delta virus infections, indicating that a higher infectious virus load alone was not driving the higher transmissibility seen in Omicron infections. Notably, no significant differences were noted in the specimen collection time frame in relation to the onset of symptoms in all groups (Figure 3D). Of note, 17.5% of samples with infectious virus were collected after 5 days from symptoms.

**Figure 3.**
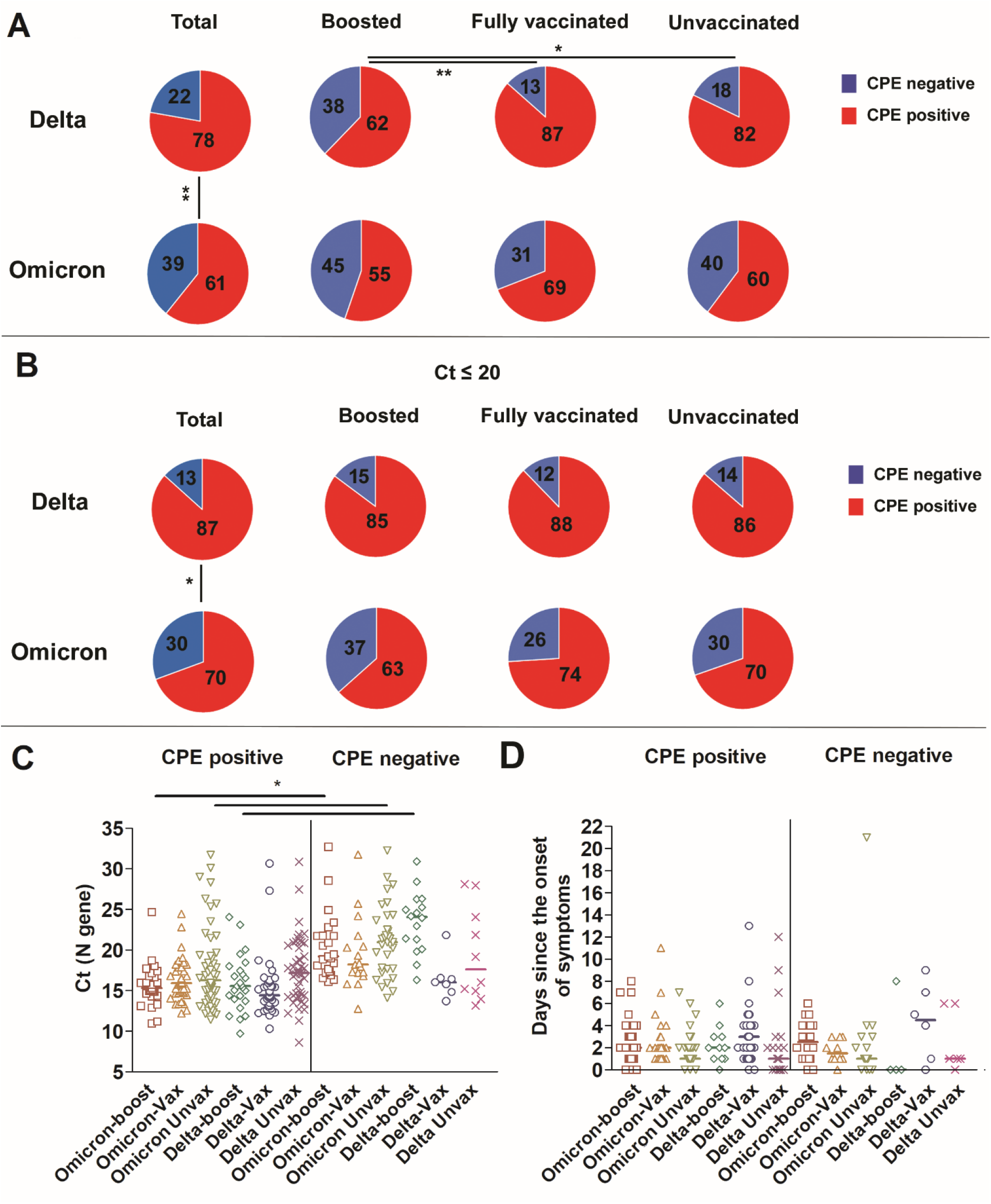
Recovery of infectious virus from respiratory samples of patients infected with Delta or Omicron. A) Percent CPE positives and negatives for Delta and Omicron; total, patients who received a booster, fully vaccinated, and unvaccinated groups. Chi-squared test * p < 0.05, ** p < 0.001. B) Percent CPE positives and negatives for Delta and Omicron; total, patients who received a booster, fully vaccinated, and unvaccinated groups with Ct values less than 20. Chi-squared test * p < 0.05. C) Ct range and medians of Delta and Omicron samples CPE positive and negative. One-way ANOVA * p < 0.05 D) Distribution of sample collection time from each group in relation to days from the onset of symptoms.

### Localized SARS-CoV-2 IgG in nasal specimens

We previously showed that SARS-CoV-2 IgG levels in the upper respiratory tract are higher in samples from vaccinated individuals and correlate with less recovery of infectious virus on cell culture (11). To compare the anti-SARS-CoV-2 IgG levels between patients who received a booster and fully vaccinated patients, ELISA was performed on upper respiratory samples from the Omicron and Delta infected groups. As expected, a significant increase in localized IgG levels was observed in patients who received a booster (Figure 4A, p < 0.0001) and IgG levels were higher in samples with no detectable infectious virus compared to those with infectious virus (Figure 4A, p < 0.0001). The anti-SARS-CoV-2 IgG levels were higher in the boosted and Omicron-infected group compared to the Omicron-infected and fully vaccinated groups but this was not significant in the Delta-infected groups (Figure 4B, p < 0.0001). Both Omicron and Delta infected patients that had infectious virus showed a significant decrease in anti-SARS-CoV-2 IgG levels when compared to specimen with no infectious virus (Figure 4C, p < 0.05). The data indicate that anti-SARS-CoV-2 antibody levels in the upper respiratory tract can be increased after booster vaccination but the presence of infectious virus with either Omicron or Delta infection is associated with lower local levels of vaccination induced antibodies.

**Figure 4.**
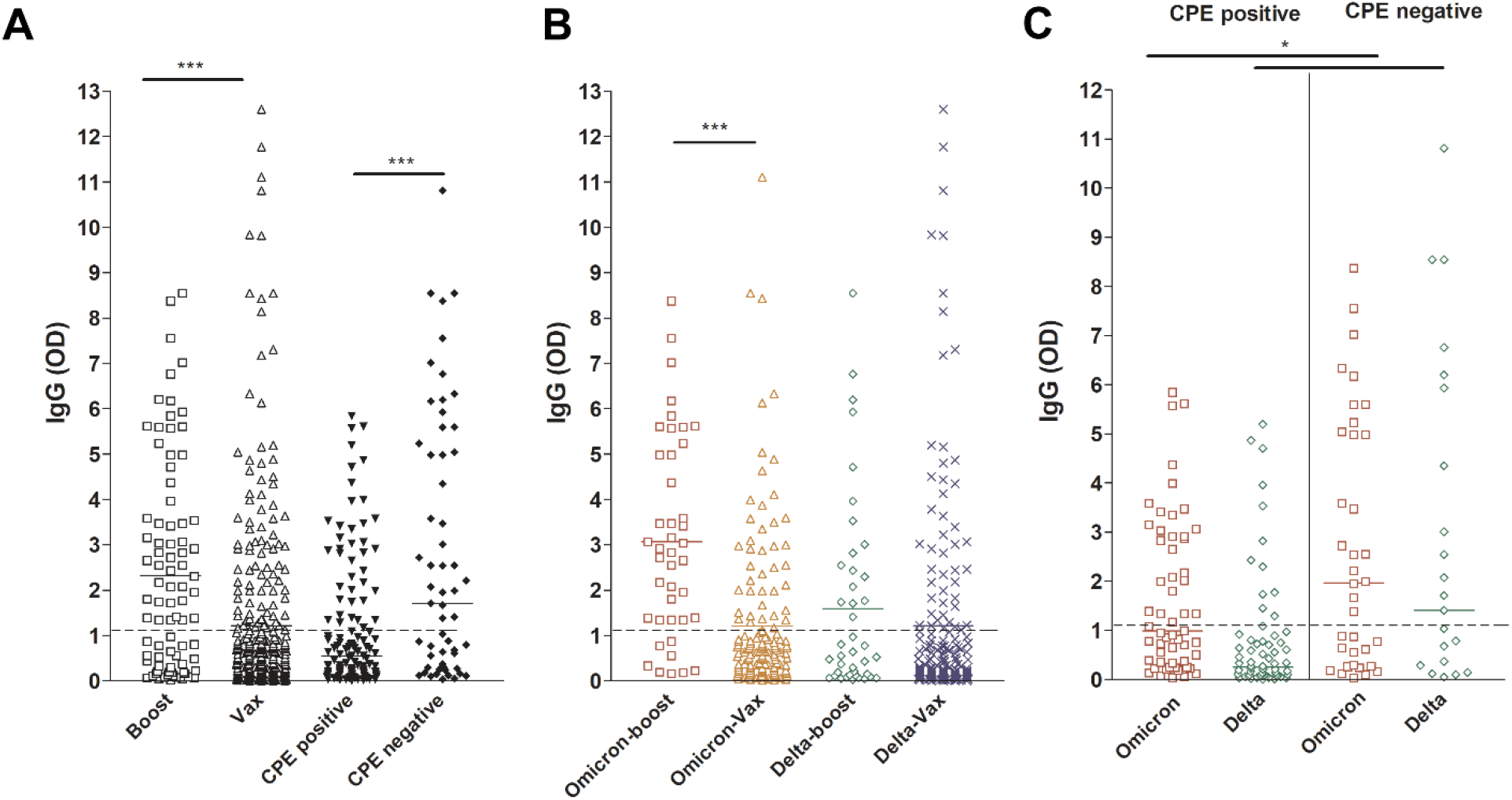
SARS-CoV-2 IgG levels in upper respiratory samples of infected vaccinated patients. Boost, patients with booster dose; Vax, fully vaccinated patients who didn’t receive a booster dose. Dashed lines demarcate the limit of borderline and negative ELISA results as specified per assay’s package insert.

## Discussion

In this study, we provide a comparison between Omicron and Delta infected patients from the transition from Delta to Omicron dominance. Using this tight time frame also controls for the timing of samples’ collection in relation to vaccinations and booster doses between the two groups in addition to the implemented community measures of infection control, including masking and social distancing. Our data showed that Omicron infected patients were associated with higher infection rates in vaccinated individuals and those who received booster vaccinations but admissions, ICU level cares, and mortality were significantly less. Samples collected from Omicron infected patients had equivalent viral loads when compared to samples collected from Delta infected individuals regardless of the vaccination status.

Recovery of infectious virus on cell culture was less frequent in the Omicron group with no significant impact of vaccination with the exception of the Delta individuals who received a booster dose, this group showed a significantly reduced recovery of infectious virus when compared to the Delta fully vaccinated and unvaccinated groups. Consistent with our previously reported observations (11), samples with successful recovery of infectious virus on cell culture correlated with less IgG levels in the respiratory samples. In this study, we also report a significant increase of IgG in samples collected from patients who received a booster dose.

The Omicron variant was first reported from South Africa early in November 2021. The first case was reported from the US on December 1^st^ 2021, a few days after the WHO classified it as a variant of concern (16). The first case we identified as a part of our SARS-CoV-2 genomic surveillance was from a patient who developed symptoms during the last week of November 2021. A very quick increase in Omicron detection correlated with a marked increase in the overall SARS-CoV-2 positivity to reach an average of close to 50% in symptomatic patients in the last week of December. Notably, the Omicron detection and rapid increase occurred during a spike in Delta circulation. In contrast, the Delta displacement of Alpha variant occurred in a time of markedly low circulation of the latter (Figure S1 and (11)). The rapid increase in Omicron cases could be explained by either an increase in overall transmissibility of this variant or due to the enhanced immune evasion of Omicron through multiple mutations in the Spike protein. In a study that compared the house hold contacts of Delta and Omicron infected patients, the secondary attack rate was higher with Omicron, particularly in fully vaccinated and boosted contacts (17). Notably, 53.1% of the Omicron infected patients from our cohort were fully vaccinated or received booster doses and there was no difference in the presence of Omicron infectious virus in either the unvaccinated, vaccinated or boosted groups, suggesting little effect of vaccination on infectious virus load. Multiple studies have shown that the neutralization of the Omicron is reduced compared to Delta or prior variants (18-21). Interestingly, there were fewer numbers of specimen containing infectious virus in the Omicron group compared to the Delta group, indicating that the presence of infectious virus alone may not explain the higher transmissibility of Omicron.

The omicron genome contains 32 amino acid changes in the spike protein, within its NTD, RBD, and close to the furin cleavage region, some of which are shared with prior variants of concern and were previously characterized (22, 23). Interestingly, those changes are expected to impact the binding to the host receptor ACE2 and alter membrane fusion (24). This likely explains the notable phenotypic changes of Omicron in cell culture and animal studies and the change in the viral tropism. Omicron was shown to cause a mild disease in animals and replicate less efficiently in the lungs (25). In addition, the use of the Vero-TMPRSS2 cell line which initially showed an enhanced sensitivity for the recovery of SARS-CoV- 2 (26), was also impacted by changes within Omicron. The slower viral growth might indicate an alternative entry pathway that is less dependent on TMPRSS2 (27). This is consistent with our observations that samples from the Omicron infected group were associated with less recovery of infectious virus on this cell line, yet, using this cell line allowed the comparisons between different groups based on their vaccination status. Our data indicates that the recovery of infectious virus with Omicron is not impacted by booster vaccination, which was not the case with Delta infected patients. Even though, when we strictly limited our analysis to samples with Ct values less than 20, the recovery of infectious virus from the Delta infected, boosted group was equivalent to other groups. IgG levels were significantly higher in samples from boosted, vaccinated patients and those with no infectious virus. Taken together, we believe that Omicron evasion of preexisting immunity contributes to lessen the impact of booster vaccination on the recovery of infectious virus, which might contribute to the increased transmission, even in individuals who receive booster vaccination.

The most recent CDC guidelines for infection control and isolation indicates that in a contingency status, health care workers can quarantine for 5 days from the onset of symptoms if asymptomatic or with mild to moderate symptoms (28). Our study showed that 17.5% of CPE positive samples were collected after 5 days from symptoms’ onset. Our data indicates that it is not uncommon to recover infectious virus after 5 days from symptoms regardless of the vaccination status when patients have symptoms. Hence care should be taken when making a release from quarantine decisions, especially when patients are showing symptoms.

The limitations of our study include the retrospective nature of data collections which doesn’t allow the collection of baseline serum and respiratory IgG levels. Antibody neutralization assays and quantification of viruses from clinical samples were not conducted as a part of this study nor were Omicron or Delta specific IgG assays. Clinical data was compiled from patients tested and admitted to the Johns Hopkins Health System. It is possible that patients who tested positive in our system then sought additional clinical care, including admission, at a different hospital not affiliated with Johns Hopkins. While this could artificially lower our admission rates, we have no reason to think that this was different in the first half of December, when Delta was predominant, compared to the end of December.

In conclusion, we show a significant reduction in disease severity with Omicron when compared to Delta, yet we show that Omicron associates with significant increases in infections of fully and booster vaccinated individuals. It is important to note that admitted patients didn’t show a significant difference in the use of supplementary oxygen, ICU level care, stressing the importance of taking infection control precautions and raising an awareness that Omicron infections should not be underestimated.

## Data Availability

 All data produced in the present study are available upon reasonable request to the authors

## Author contributions

AF, RE, JS. data collection and data interpretation. CPM. data collection and analysis OA, JMN, NG, NS, ML, MF, DCG. data acquisition and collection. AP. cell culture, scientific and manuscript revision. EK. clinical data collection and scientific and manuscript revision. HHM. study design, data collection and analysis, data interpretation, writing, fund acquisition.

## Data sharing

Whole genome data were made available publicly and raw genomic data requests could be directed to HHM.

## Acknowledgement

This study was only possible with the unique efforts of the Johns Hopkins Clinical Microbiology Laboratory faculty and staff. HHM is supported by the HIV Prevention Trials Network (HPTN) sponsored by the National Institute of Allergy and Infectious Diseases (NIAID). Funding was provided by the Johns Hopkins Center of Excellence in Influenza Research and Surveillance (HHSN272201400007C), National Institute on Drug Abuse, National Institute of Mental Health, and Office of AIDS Research, of the NIH, DHHS (UM1 AI068613), the NIH RADx-Tech program (3U54HL143541-02S2), National Institute of Health RADx-UP initiative (Grant R01 DA045556-04S1), Centers for Disease Control (contract 75D30121C11061), the Johns Hopkins University President’s Fund Research Response, the Johns Hopkins department of Pathology, and the Maryland department of health. EK was supported by Centers for Disease Control and Prevention (CDC) MInD-Healthcare Program (Grant Number U01CK000589). The views expressed in this manuscript are those of the authors and do not necessarily represent the views of the National Institute of Biomedical Imaging and Bioengineering; the National Heart, Lung, and Blood Institute; the National Institutes of Health, or the U.S. Department of Health and Human Services.

---

**Figure S1.**
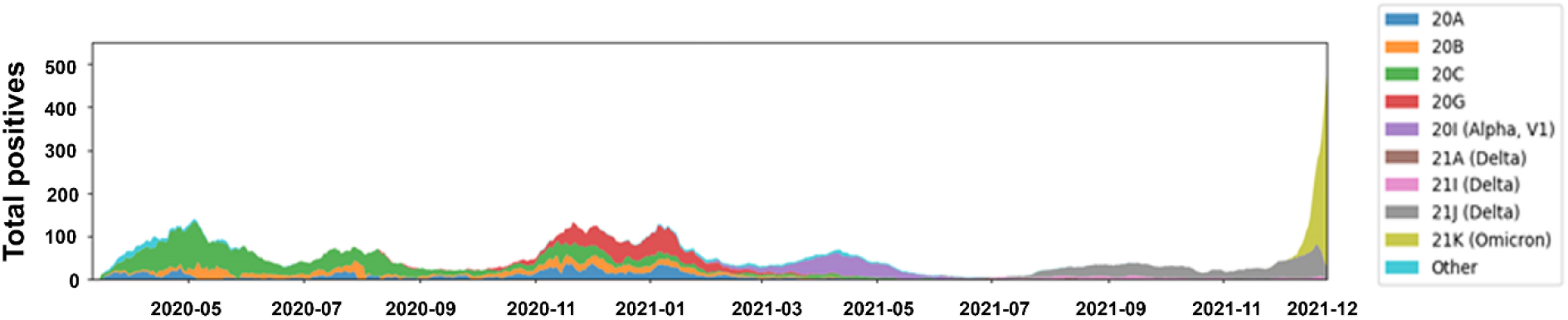
SARS-CoV-2 positivity and variants trends March 2020- December 2021. SARS-CoV-2 clade distribution between March 2020 and December 2021 relative to the 7 day rolling average positives from Johns Hopkins system.

**Figure S2.**
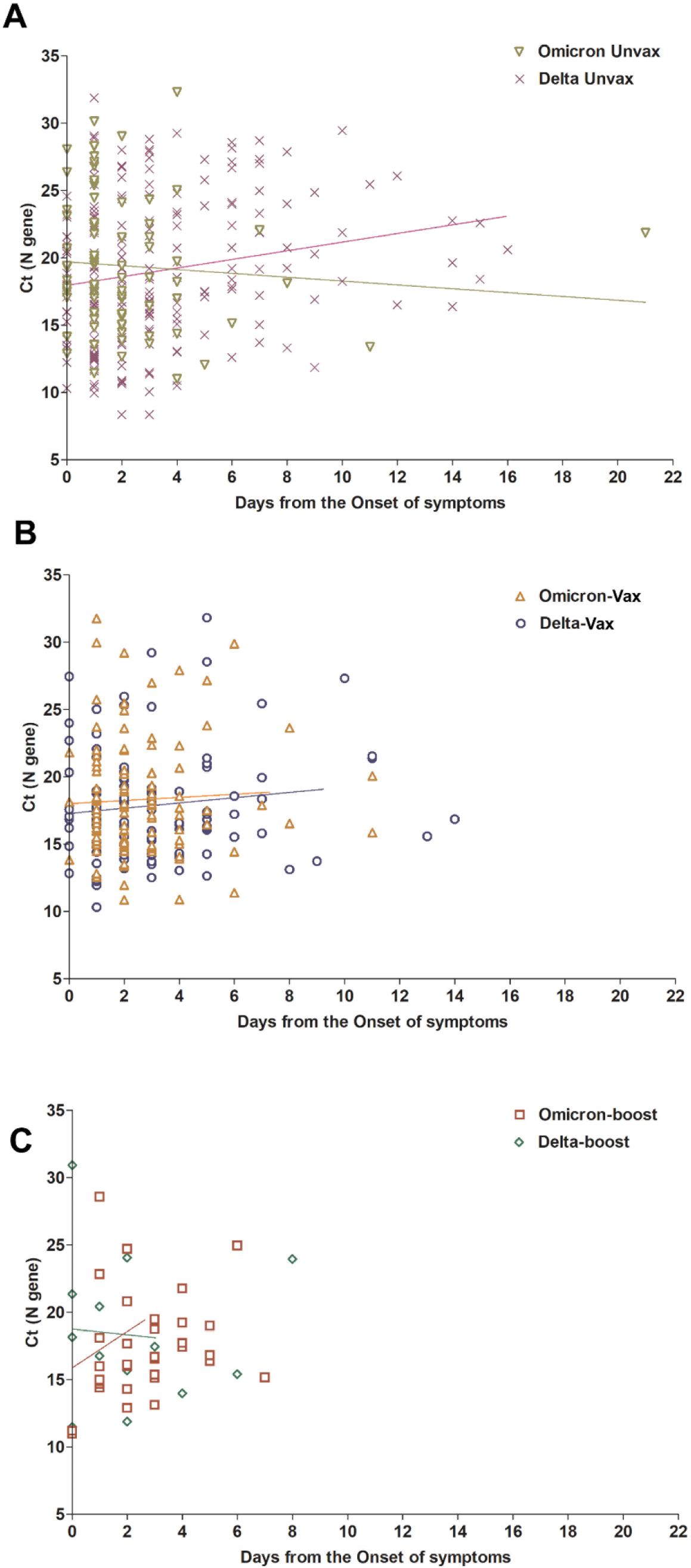
Omicron and Delta variants cycle threshold (Ct) values in upper respiratory samples. Ct values of Omicron and Delta variants broken down by vaccination status and associated with days after the onset of symptoms in A) Unvax, unvaccinated, B) Vax, fully vaccinated patients who didn’t receive a booster, and C) boost, patients with booster vaccinations.

**Table S1.** Clinical and metadata of Delta and Omicron vaccinated and unvaccinated patients. Statistics for ages were calculated by t test and all other statistics were calculated by Chi-squared test. stdev; standard deviation.

|  | Omicron |  |  |  |  |  |  | Delta |  |  |  |  |  |  |  |
| --- | --- | --- | --- | --- | --- | --- | --- | --- | --- | --- | --- | --- | --- | --- | --- |
|  | Fully Vaccinated |  | Boosted |  | Unvaccinated |  | p (boosted to unvaccinated, Omicron) | Fully Vaccinated |  | Boosted |  | Unvaccinated |  | p (boosted to unvaccinated, Delta) | P (boosted Omicron to boosted Delta) |
| Total | 461 | % | 134 | % | 488 | % |  | 282 | % | 83 | % | 518 | % |  |  |
| Symptomatic | 420 | 91.1 | 127 | 94.8 | 454 | 93.0 |  | 255 | 90.4 | 74 | 89.2 | 385 | 74.3 |  |  |
| Asymptomatic | 41 | 8.9 | 7 | 5.2 | 34 | 7.0 |  | 27 | 9.6 | 9 | 10.8 | 32 | 6.2 |  |  |
| Age (median) | 32 (17.9) |  | 34 (19.3) |  | 31.8 (18.8) |  |  | 36 (22.6) |  | 39 (25.1) |  | 34 (23.6) |  |  |  |
| Comorbidities |  |  |  |  |  |  |  |  |  |  |  |  |  |  |  |
| Hypertension | 95 | 20.6 | 32 | 23.9 | 109 | 22.3 | 0.7 | 98 | 34.8 | 28 | 33.7 | 149 | 28.8 | 0.3 | 0.12 |
| Pregnancy | 37 | 8.0 | 13 | 9.7 | 30 | 6.1 | 0.2 | 21 | 7.4 | 3 | 3.6 | 27 | 5.2 | 0.8 | 0.114 |
| Lung Disease | 85 | 18.4 | 22 | 16.4 | 111 | 22.7 | 0.1 | 62 | 22.0 | 22 | 26.5 | 115 | 22.2 | 0.4 | 0.08 |
| Kidney Disease | 30 | 6.5 | 6 | 4.5 | 30 | 6.1 | 0.5 | 34 | 12.1 | 16 | 19.3 | 72 | 13.9 | 0.2 | 0.0008 |
| Immunosuppression | 51 | 11.1 | 15 | 11.2 | 51 | 10.5 | 0.9 | 45 | 16.0 | 20 | 24.1 | 86 | 16.6 | 0.1 | 0.01 |
| Diabetes | 45 | 9.8 | 15 | 11.2 | 43 | 8.8 | 0.4 | 43 | 15.2 | 17 | 20.5 | 74 | 14.3 | 0.1 | 0.07 |
| Heart Failure | 14 | 3.0 | 1 | 0.7 | 13 | 2.7 | 0.3 | 24 | 8.5 | 6 | 7.2 | 30 | 5.8 | 0.6 | 0.01 |
| Atrial Fibrillation | 7 | 1.5 | 2 | 1.5 | 8 | 1.6 | 1.0 | 12 | 4.3 | 6 | 7.2 | 27 | 5.2 | 0.4 | 0.01 |
| Smoker | 54 | 11.7 | 10 | 7.5 | 45 | 9.2 | 0.6 | 42 | 14.9 | 11 | 13.3 | 84 | 16.2 | 0.6 | 0.0002 |
| Cerebrovascular Disease | 20 | 4.3 | 4 | 3.0 | 20 | 4.1 | 0.8 | 21 | 7.4 | 5 | 6.0 | 41 | 7.9 | 0.7 | 0.3 |
| Cancer | 104 | 22.6 | 38 | 28.4 | 103 | 21.1 | 0.1 | 65 | 23.0 | 26 | 31.3 | 112 | 21.6 | 0.1 | 0.6 |
| Coronary Artery Disease | 41 | 8.9 | 10 | 7.5 | 31 | 6.4 | 0.7 | 45 | 16.0 | 15 | 18.1 | 88 | 17.0 | 0.8 | 0.03 |
| Outcome |  |  |  |  |  |  |  |  |  |  |  |  |  |  |  |
| COVID Related Admission | 20 | 4.3 | 3 | 2.2 | 9 | 1.8 | 0.7 | 30 | 10.6 | 13 | 15.7 | 80 | 15.4 | 1.0 | 0.0007 |
| ICU Level Care | 2 | 0.4 | 1 | 0.7 | 2 | 0.4 | 0.5 | 6 | 2.1 | 2 | 2.4 | 24 | 4.6 | 0.6 | 0.6 |
| Died | 0 | 0.0 | 0 | 0.0 | 1 | 0.2 | 1.0 | 2 | 0.7 | 2 | 2.4 | 6 | 1.2 | 0.3 | 0.1 |

## References

1. Team CC-R. 2021. SARS-CoV-2 B.1.1.529 (Omicron) Variant - United States, December 1-8, 2021. MMWR Morb Mortal Wkly Rep 70:1731–1734.

2. Kannan S, Shaik Syed Ali P, Sheeza A. 2021. Omicron (B.1.1.529) - variant of concern - molecular profile and epidemiology: a mini review. Eur Rev Med Pharmacol Sci 25:8019–8022.

3. WHO. World Health Organization. Classification of Omicron (B.1.1.529): SARS-CoV-2 variant of concern. Geneva, Switzerland: World Health Organization; 2021. Accessed December 3, 2021.

4. Wolter N, Jassat W, Walaza S, Welch R, Moultrie H, Groome M, Amoako DG, Everatt J, Bhiman JN, Scheepers C, Tebeila N, Chiwandire N, du Plessis M, Govender N, Ismail A, Glass A, Mlisana K, Stevens W, Treurnicht FK, Makatini Z, Hsiao NY, Parboosing R, Wadula J, Hussey H, Davies MA, Boulle A, von Gottberg A, Cohen C. 2022. Early assessment of the clinical severity of the SARS-CoV-2 omicron variant in South Africa: a data linkage study. Lancet doi:10.1016/s0140-6736(22)00017-4.

5. Maslo C, Friedland R, Toubkin M, Laubscher A, Akaloo T, Kama B. 2021. Characteristics and Outcomes of Hospitalized Patients in South Africa During the COVID-19 Omicron Wave Compared With Previous Waves. JAMA doi:10.1001/jama.2021.24868.

6. Anonymous. Investigation of SARS-CoV-2 variants: technical briefings. UK Health Security Agency. SARS-CoV-2 variants of concern and variants under investigation in England Technical briefing 34. 14 January 2022.

7. Jarrett J, Uhteg K, Forman MS, Hanlon A, Vargas C, Carroll KC, Valsamakis A, Mostafa HH. 2021. Clinical performance of the GenMark Dx ePlex respiratory pathogen panels for upper and lower respiratory tract infections. J Clin Virol 135:104737.

8. Mostafa HH, Carroll KC, Hicken R, Berry GJ, Manji R, Smith E, Rakeman JL, Fowler RC, Leelawong M, Butler-Wu SM, Quintero D, Umali-Wilcox M, Kwiatkowski RW, Persing DH, Weir F, Loeffelholz MJ. 2020. Multi-center Evaluation of the Cepheid Xpert(R) Xpress SARS-CoV-2/Flu/RSV Test. J Clin Microbiol doi:10.1128/JCM.02955-20.

9. Mostafa HH, Hardick J, Morehead E, Miller JA, Gaydos CA, Manabe YC. 2020. Comparison of the analytical sensitivity of seven commonly used commercial SARS-CoV-2 automated molecular assays. J Clin Virol 130:104578.

10. Uhteg K, Jarrett J, Richards M, Howard C, Morehead E, Geahr M, Gluck L, Hanlon A, Ellis B, Kaur H, Simner P, Carroll KC, Mostafa HH. 2020. Comparing the analytical performance of three SARS- CoV-2 molecular diagnostic assays. J Clin Virol 127:104384.

11. Luo CH, Morris CP, Sachithanandham J, Amadi A, Gaston DC, Li M, Swanson NJ, Schwartz M, Klein EY, Pekosz A, Mostafa HH. 2021. Infection with the SARS-CoV-2 Delta Variant is Associated with Higher Recovery of Infectious Virus Compared to the Alpha Variant in both Unvaccinated and Vaccinated Individuals. Clin Infect Dis doi:10.1093/cid/ciab986.

12. Thielen PM, Wohl S, Mehoke T, Ramakrishnan S, Kirsche M, Falade-Nwulia O, Trovao NS, Ernlund A, Howser C, Sadowski N, Morris CP, Hopkins M, Schwartz M, Fan Y, Gniazdowski V, Lessler J, Sauer L, Schatz MC, Evans JD, Ray SC, Timp W, Mostafa HH. 2021. Genomic diversity of SARS-CoV-2 during early introduction into the Baltimore-Washington metropolitan area. JCI Insight 6.

13. Morris CP, Luo CH, Amadi A, Schwartz M, Gallagher N, Ray SC, Pekosz A, Mostafa HH. 2021. An Update on SARS-CoV-2 Diversity in the United States National Capital Region: Evolution of Novel and Variants of Concern. Clin Infect Dis doi:10.1093/cid/ciab636.

14. Gniazdowski V, Morris CP, Wohl S, Mehoke T, Ramakrishnan S, Thielen P, Powell H, Smith B, Armstrong DT, Herrera M, Reifsnyder C, Sevdali M, Carroll KC, Pekosz A, Mostafa HH. 2020. Repeat COVID-19 Molecular Testing: Correlation of SARS-CoV-2 Culture with Molecular Assays and Cycle Thresholds. Clin Infect Dis doi:10.1093/cid/ciaa1616.

15. Uhteg K, Amadi A, Forman M, Mostafa HH. 2021. Circulation of Non- SARS-CoV-2 Respiratory Pathogens and Coinfection with SARS-CoV-2 Amid the COVID-19 Pandemic. Open Forum Infectious Diseases doi:10.1093/ofid/ofab618.

16. CDC. 2021. Omicron Variant: What You Need to Know. Centers for Disease Control and Prevention. Last accessed January 19, 2022.

17. Lyngse FP, Mortensen LH, Denwood MJ, Christiansen LE, Møller CH, Skov RL, Spiess K, Fomsgaard A, Lassaunière MM, Rasmussen M, Stegger M, Nielsen C, Sieber RN, Cohen AS, Møller FT, Overvad M, Mølbak K, Krause TG, Kirkeby CT. 2021. SARS-CoV-2 Omicron VOC Transmission in Danish Households. medRxiv doi:10.1101/2021.12.27.21268278:2021.12.27.21268278.

18. Basile K, Rockett RJ, McPhie K, Fennell M, Johnson-Mackinnon J, Agius JE, Fong W, Rahman H, Ko D, Donavan L, Hueston L, Lam C, Arnott A, Chen SC-A, Maddocks S, O’Sullivan MV, Dwyer DE, Sintchenko V, Kok J. 2021. Improved neutralization of the SARS-CoV-2 Omicron variant after Pfizer-BioNTech BNT162b2 COVID-19 vaccine boosting. bioRxiv doi:10.1101/2021.12.12.472252:2021.12.12.472252.

19. Planas D, Saunders N, Maes P, Guivel-Benhassine F, Planchais C, Buchrieser J, Bolland W-H, Porrot F, Staropoli I, Lemoine F, Péré H, Veyer D, Puech J, Rodary J, Baele G, Dellicour S, Raymenants J, Gorissen S, Geenen C, Vanmechelen B, Wawina -Bokalanga T, Martí-Carreras J, Cuypers L, Sève A, Hocqueloux L, Prazuck T, Rey F, Simon-Loriere E, Bruel T, Mouquet H, André E, Schwartz O. 2021. Considerable escape of SARS-CoV-2 Omicron to antibody neutralization. Nature doi:10.1038/s41586-021-04389-z.

20. Schmidt F, Muecksch F, Weisblum Y, Da Silva J, Bednarski E, Cho A, Wang Z, Gaebler C, Caskey M, Nussenzweig MC, Hatziioannou T, Bieniasz PD. 2021. Plasma Neutralization of the SARS-CoV- 2 Omicron Variant. New England Journal of Medicine doi:10.1056/NEJMc2119641.

21. Rössler A, Riepler L, Bante D, Laer Dv, Kimpel J. 2021. SARS-CoV-2 B.1.1.529 variant (Omicron) evades neutralization by sera from vaccinated and convalescent individuals. medRxiv doi:10.1101/2021.12.08.21267491:2021.12.08.21267491.

22. Starr TN, Greaney AJ, Addetia A, Hannon WW, Choudhary MC, Dingens AS, Li JZ, Bloom JD. 2021. Prospective mapping of viral mutations that escape antibodies used to treat COVID-19. Science 371:850–854.

23. Plante JA, Mitchell BM, Plante KS, Debbink K, Weaver SC, Menachery VD. 2021. The variant gambit: COVID-19’s next move. Cell Host Microbe 29:508–515.

24. Zhang J, Cai Y, Lavine CL, Peng H, Zhu H, Anand K, Tong P, Gautam A, Mayer ML, Rits-Volloch S, Wang S, Sliz P, Wesemann DR, Yang W, Seaman MS, Lu J, Xiao T, Chen B. 2022. Structural and functional impact by SARS-CoV-2 Omicron spike mutations. bioRxiv doi:10.1101/2022.01.11.475922:2022.01.11.475922.

25. Diamond M, Halfmann P, Maemura T, Iwatsuki-Horimoto K, Iida S, Kiso M, Scheaffer S, Darling T, Joshi A, Loeber S, Foster S, Ying B, Whitener B, Floyd K, Ujie M, Nakajima N, Ito M, Wright R, Uraki R, Li R, Sakai Y, Liu Y, Larson D, Osorio J, Hernandez-Ortiz J, ÄŒiuoderis K, Florek K, Patel M, Bateman A, Odle A, Wong LY, Wang Z, Edara VV, Chong Z, Thackray L, Ueki H, Yamayoshi S, Imai M, Perlman S, Webby R, Seder R, Suthar M, Garcia-Sastre A, Schotsaert M, Suzuki T, Boon A, Kawaoka Y, Douek D, Moliva J, Sullivan N, et al. 2021. The SARS-CoV-2 B.1.1.529 Omicron virus causes attenuated infection and disease in mice and hamsters. Res Sq doi:10.21203/rs.3.rs-1211792/v1.

26. Matsuyama S, Nao N, Shirato K, Kawase M, Saito S, Takayama I, Nagata N, Sekizuka T, Katoh H, Kato F, Sakata M, Tahara M, Kutsuna S, Ohmagari N, Kuroda M, Suzuki T, Kageyama T, Takeda M. 2020. Enhanced isolation of SARS-CoV-2 by TMPRSS2-expressing cells. Proceedings of the National Academy of Sciences 117:7001–7003.

27. Zhao H, Lu L, Peng Z, Chen LL, Meng X, Zhang C, Ip JD, Chan WM, Chu AW, Chan KH, Jin DY, Chen H, Yuen KY, To KK. 2022. SARS-CoV-2 Omicron variant shows less efficient replication and fusion activity when compared with Delta variant in TMPRSS2-expressed cells. Emerg Microbes Infect 11:277–283.

28. CDC. 2021. Interim Guidance for Managing Healthcare Personnel with SARS-CoV-2 Infection or Exposure to SARS-CoV-2. Centers for Disease Control and Prevention. Last accessed 1/20/22.

